# Shared Genetic Architecture Between Endometriosis and Psychiatric Conditions May Explain Comorbidity

**DOI:** 10.1101/2025.10.22.25338556

**Authors:** Marika Rostvall, Cecilia Magnusson, Kristina Gemzell-Danielsson, Johanna Sieurin, Anders D. Børglum, Jakob Grove, Mette Nyegaard, Anxiety Disorder Working Group of the Psychiatric Genomics Consortium, Autism Spectrum Disorder Working Group of the Psychiatric Genomics Consortium, Emma Bränn, Kyriaki Kosidou, Jacob Bergstedt, Viktor H. Ahlqvist

## Abstract

Endometriosis is a common chronic gynecological disorder with complex and poorly understood etiology and no definitive cure. Beyond pain and infertility, affected individuals frequently experience psychiatric comorbidities, often attributed to the burden of chronic symptoms. Emerging evidence suggests this explanation is incomplete, and shared biological mechanisms may also contribute. Here, we integrate large-scale genomic data to characterize the genetic overlap between endometriosis and a broad spectrum of psychiatric conditions. We found no evidence that genetic liability to endometriosis increases the risk of psychiatric conditions. In contrast, genetic liability to psychiatric conditions, particularly major depressive disorder and related traits, was associated with increasing the risk of endometriosis. Polygenic analyses revealed extensive shared genetic architecture, with nearly all variants influencing endometriosis also implicated in depression. Leveraging this overlap in a multivariate GWAS, we identify 606 independent genome-wide significant variants contributing to shared liability and implicate convergent biological pathways - particularly brain-related mechanisms - providing a foundation for mechanistic studies and potential strategies to mitigate psychiatric comorbidity in endometriosis.

## Introduction

Endometriosis is a chronic, estrogen-dependent inflammatory disorder characterized by the growth of endometrial-like tissue outside the uterus, most commonly within the pelvic cavity. Affecting ∼10% of women of reproductive age, endometriosis remains underdiagnosed and poorly understood(1). The estimated heritability of endometriosis is around 50% (2, 3), with an estimated 26% due to common genetic variants(4). Symptoms include dysmenorrhea, dyspareunia, chronic pelvic pain, and endometriosis is a leading cause of female subfertility(1). Despite the high prevalence and profound impact on quality of life, research into etiology has lagged, diagnosis is often delayed, and treatment options remain limited.

Beyond pain and reproductive morbidity, many population studies have reported elevated rates of depression and anxiety among affected individuals(5–7). While often attributed to the burden of chronic pain, emerging data suggest the relationship may not be purely secondary. Notably, an increased incidence of bipolar disorder, ADHD, and personality disorders—where the pain link might be less clear—has been observed(6), indicating potential involvement of shared underlying biological mechanisms. Candidate pathways include systemic inflammation and immune dysregulation, implicated in the pathogenesis of endometriosis and psychiatric conditions(8, 9), and genetic or environmental predispositions to heightened pain sensitivity or psychological distress (10, 11).

Recent genetic studies partly support a shared genetic liability, reporting moderate genetic correlations between endometriosis and major depressive disorder (MDD), anxiety, eating disorders, and post-traumatic stress disorder (PTSD)(12–14). Mendelian randomization studies also suggest that liability to mood-related traits increase endometriosis risk(12, 14–17). However, most prior work has been constrained by limited statistical power, due to the small size of prior endometriosis GWAS’s, and therefore could not reliably resolve causal directionality or disentangle shared mechanisms. More granular and better-powered analyses are needed to clarify whether co-occurrence reflects causal pathways, shared biology, or both. The latest GWAS, identifying 42 genome-wide significant loci across 49 signals and explaining up to 5% of disease variance (18), now provides an unprecedented opportunity to revisit these questions.

In this study, we leverage this large-scale genetic data together with similar efforts in psychiatric genomics to gain deeper insight into why individuals with endometriosis exhibit such high rates of psychiatric comorbidity, and to determine whether this may reveal clues to shared etiology and opportunities for prevention or treatment.

## Results

We used the largest available GWAS of endometriosis(18), comprising over 20,000 cases and 400,000 controls of European ancestry, together with large-scale psychiatric GWAS from the Psychiatric Genomics Consortium and other consortia(19–28) (**Figure 1**). After harmonizing these large-scale results, we (i) estimated global genetic correlations between endometriosis and 11 psychiatric conditions and 11 depressive symptoms dimensions; (ii) mapped local correlations to identify regional hotspots of shared architecture; (iii) assessed causal directionality using bidirectional Mendelian randomization; (iv) decomposed the polygenicity of shared genetic variants, and (v) since we find evidence of strong genetic overlap between endometriosis and depression-related traits, we applied genomic structural equation modelling to perform a multivariate GWAS of endometriosis and MDD and tissue expression analysis to examine in which human tissues the overlapping genetic liability is expressed.

**Figure 1:**
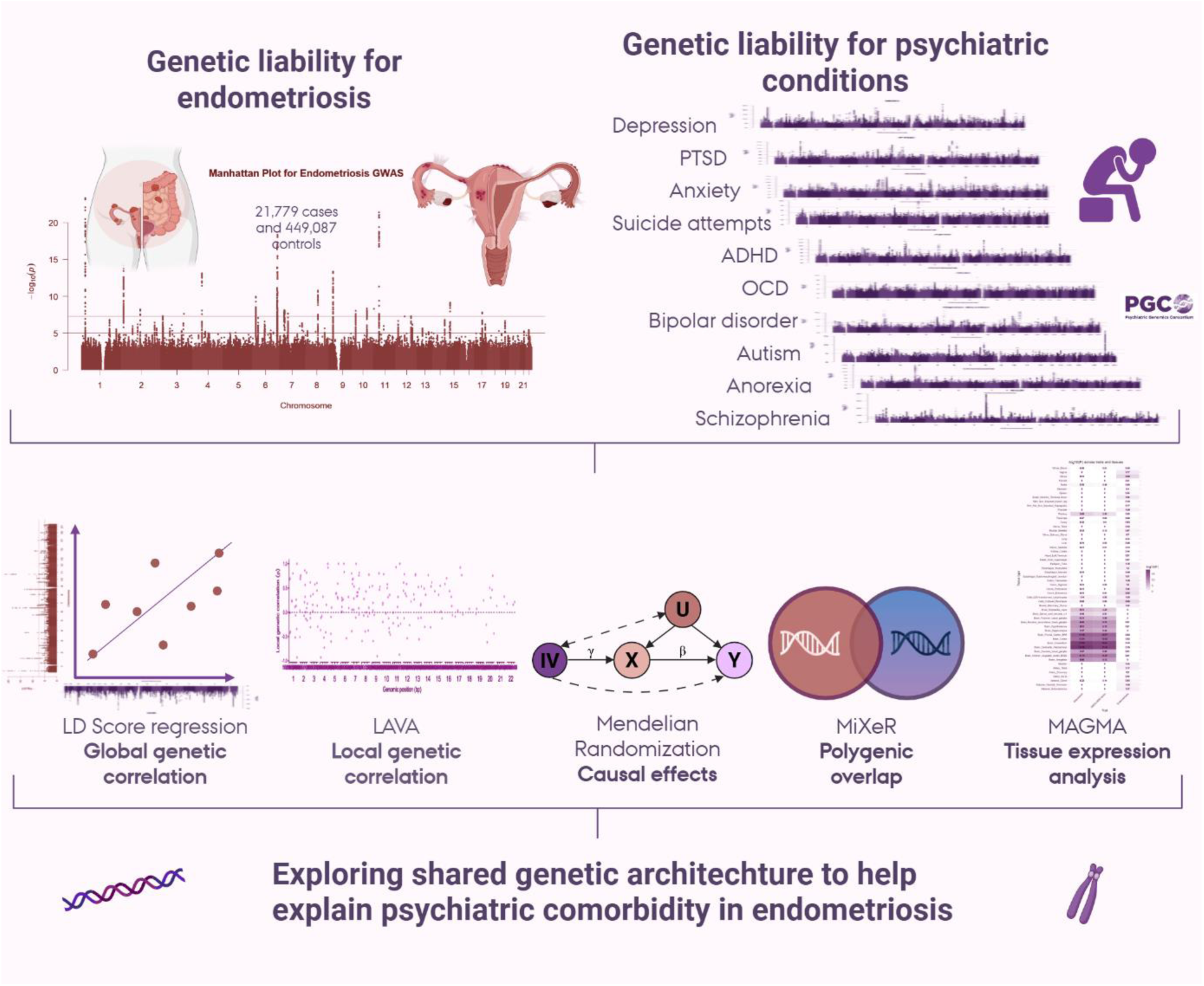
Overview of methods and data used.

### Global genetic correlation between endometriosis and psychiatric conditions and depressive symptoms

Using linkage disequilibrium score regression (LDSC)(29), we observed moderate to strong positive genome-wide genetic correlations between endometriosis and several psychiatric conditions (**Figure 2A**, supplementary table 2). The strongest correlations were observed with depression (global genetic correlation [r_g_] = 0.36, P-value *[P]* = 1.75 × 10⁻³⁰), PTSD (r_g_ = 0.34, *P* = 5.12 × 10⁻¹⁷), anxiety disorder (r_g_ = 0.29, *P* = 5.73 × 10⁻¹⁶), suicide attempts (r_g_ = 0.26, *P* = 1.91 × 10⁻⁵), and attention-deficit/hyperactivity disorder (ADHD) (r_g_ = 0.21, *P* =8.09 × 10⁻⁶). Genetic correlations with obsessive-compulsive disorder (OCD) and bipolar disorder were weaker (r_g_ = 0.13 for both) but still significant. All nominally statistically significant (*P*>0.05) global genetic correlations survived false discovery rate (FDR) correction for multiple testing. There were no global genetic correlations with anorexia, autism, or schizophrenia.

**Figure 2.**
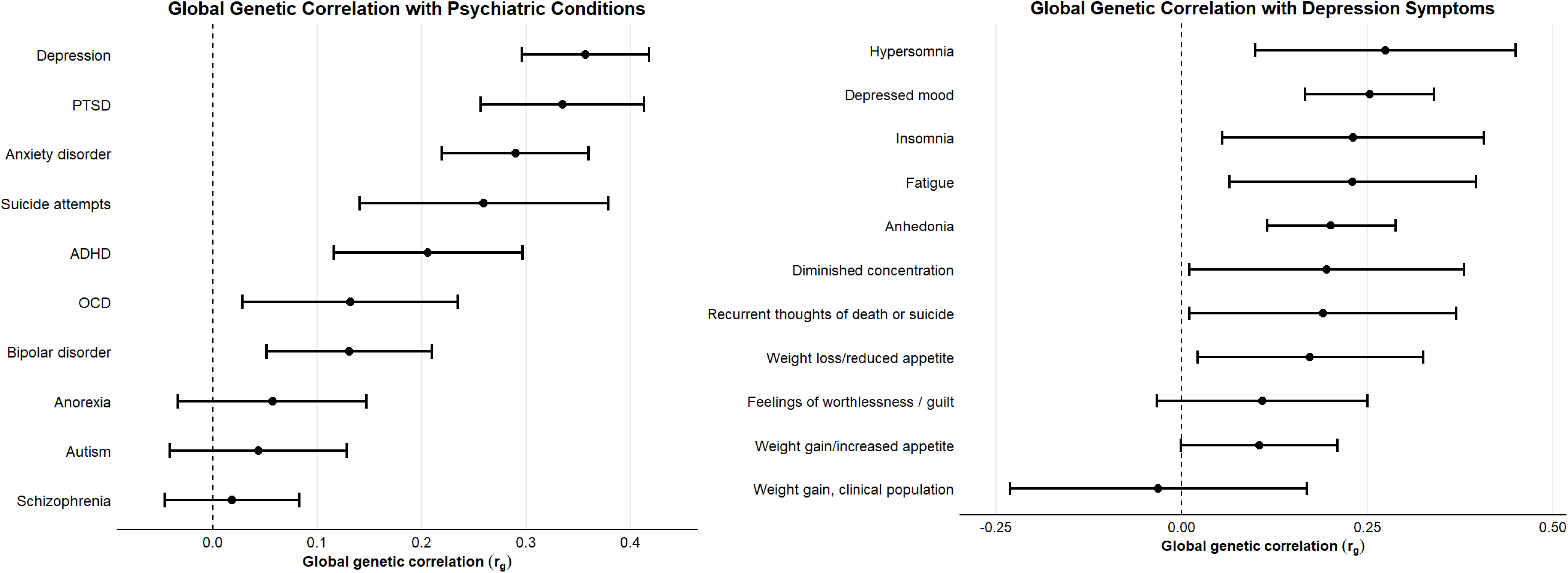
Globalgeneticcorrelation (r_g_) betweenendometriosisand differentpsychiatricconditions(A) and depressive symptoms(B). Estimated using LDSC. Points show r_g_estimates, with horizontal lines indicating 95% confidence intervals.

To further explore the apparent depressive signature in the pattern of global genetic correlations, we examined global genetic correlations between endometriosis and specific depressive symptoms (**Figure 2B**, supplementary table 3). We observed statistically significant positive correlations for a broad range of symptoms, including hypersomnia (r_g_ = 0.28, *P* = 2.00 × 10⁻³), depressed mood (r_g_ = 0.25, *P* = 1.12 × 10⁻⁸), insomnia (r_g_ = 0.23, *P* = 0.01), fatigue (rg = 0.23, *P* = 0.01), anhedonia (r_g_ = 0.20, *P* = 4.70 × 10⁻⁶), diminished concentration (r_g_ = 0.20, *P* = 0.04), recurrent thoughts of death or suicide (r_g_ = 0.19, *P* =0.04), and weight loss or reduced appetite (r_g_ = 0.17, *P* = 0.03). All nominally statistically significant (*P*>0.05) correlated symptoms, except diminished concentration and recurrent thoughts of death or suicide, survived FDR correction. Overall, no single symptom exhibited a markedly stronger genetic correlation than the others, suggesting that the genetic link between endometriosis and depression is not driven by a specific subset of depressive symptoms but may instead reflect a broad pattern of shared genetic architecture.

### Local genetic correlation between endometriosis and psychiatric conditions

We used Local Analysis of [co]Variant Association (LAVA)(30) to estimate local genetic correlations at 2,495 loci across the genome. We identified the largest number of statistically significant local genetic correlations with depression (n loci = 40, with local r_g_ ranging from -0.931 to 1), of which 88% showed positive correlation with endometriosis (**Figure 3**). None of these signals survived phenotype-by-phenotype FDR correction, potentially suggesting that the genetic overlap is polygenic and broadly distributed, rather than driven by a small number of loci exhibiting strong local genetic correlations. A similar pattern was observed for anxiety and bipolar disorder, each with 17 loci with nominally statistically significant local genetic correlations, 71% of which showed positive correlations. For PTSD, 17 loci were also statistically significant, though only 58% were in a positive direction.

**Figure 3.**
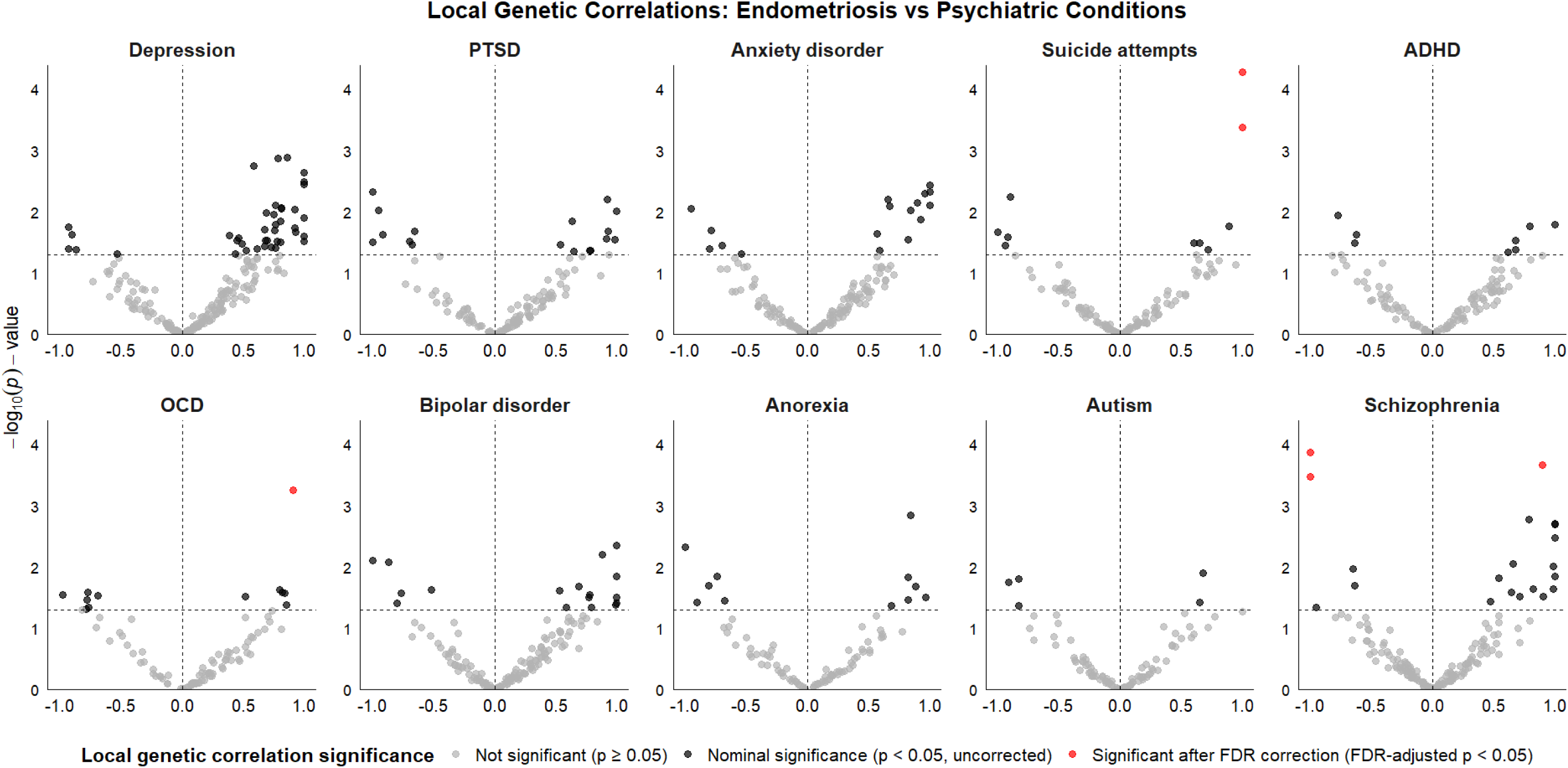
Localgeneticcorrelationsbetweenendometriosisandpsychiatricconditions. Localcorrelationswereestimatedusing LAVAacross 2,495 genomic loci. Each point represents the local genetic correlation coefficient (r_g_); light grey indicates no signal, black nominal associations (p < 0.05), and red loci significant after FDR correction.

Despite a low genome-wide genetic correlation estimated using LDSC (r_g_ = 0.02, p-value = 0.58), schizophrenia showed 16 nominally statistically significant loci, including one positive correlation that survived FDR correction (local r_g_ = 0.901, Q-value = 0.02, hg19 build; chr13:101,573,737-103,050,417) (**Figure 3**). This region encompasses *PIK3R1*, a regulator of PI3K/AKT signalling, with somatic mutations common in endometrial cancer and variation in this pathway implicated in schizophrenia, bipolar disorder, and neuroinflammation in major depressive disorder(31–34). However, several loci exhibited strong negative local genetic correlations between schizophrenia and endometriosis, potentially contributing to the low global correlation. For suicide attempts, 12 loci were statistically significant, including two positive correlations that survived stringent FDR correction (local r_g_ = 1.0, Q-value = 0.02, chr6:143,083,348-144,791,455 and local r_g_ = 1.0, Q-value = 0.01, chr7:46,247,336-47,253,454). The chr6 locus spans ESR1 and SOX4, genes involved in estrogen signaling and neurodevelopment(35, 36), while the chr7 locus includes IKZF1 and TRIM24, regulators of immune function and hormone-responsive transcription(37, 38), together highlighting pathways relevant to both endometriosis and psychiatric risk. Anorexia, ADHD, and OCD had relatively few bivariate local signals with endometriosis, albeit with a slight predominance of positive correlations. Autism had the fewest loci with statistically significant local genetic correlations with endometriosis, with two positive and three negative correlations.

### Bidirectional causal effects

Using Mendelian randomization(39) to examine bidirectional causal effects, we found no evidence that genetic liability for endometriosis confers increased risk for most of the evaluated psychiatric conditions (**Figure 4a**, Supplementary Table 5). For example, the results did not support an increased risk of depression exceeding 3% (OR = 1.01, 95% Confidence Interval [CI]: 0.98–1.03). The exception was anorexia nervosa, where an inverse association was observed using the IVW estimator (OR = 0.92, 95% CI: 0.86–0.99); however, this finding was no longer statistically significant after FDR-correction.

**Figure 4.**
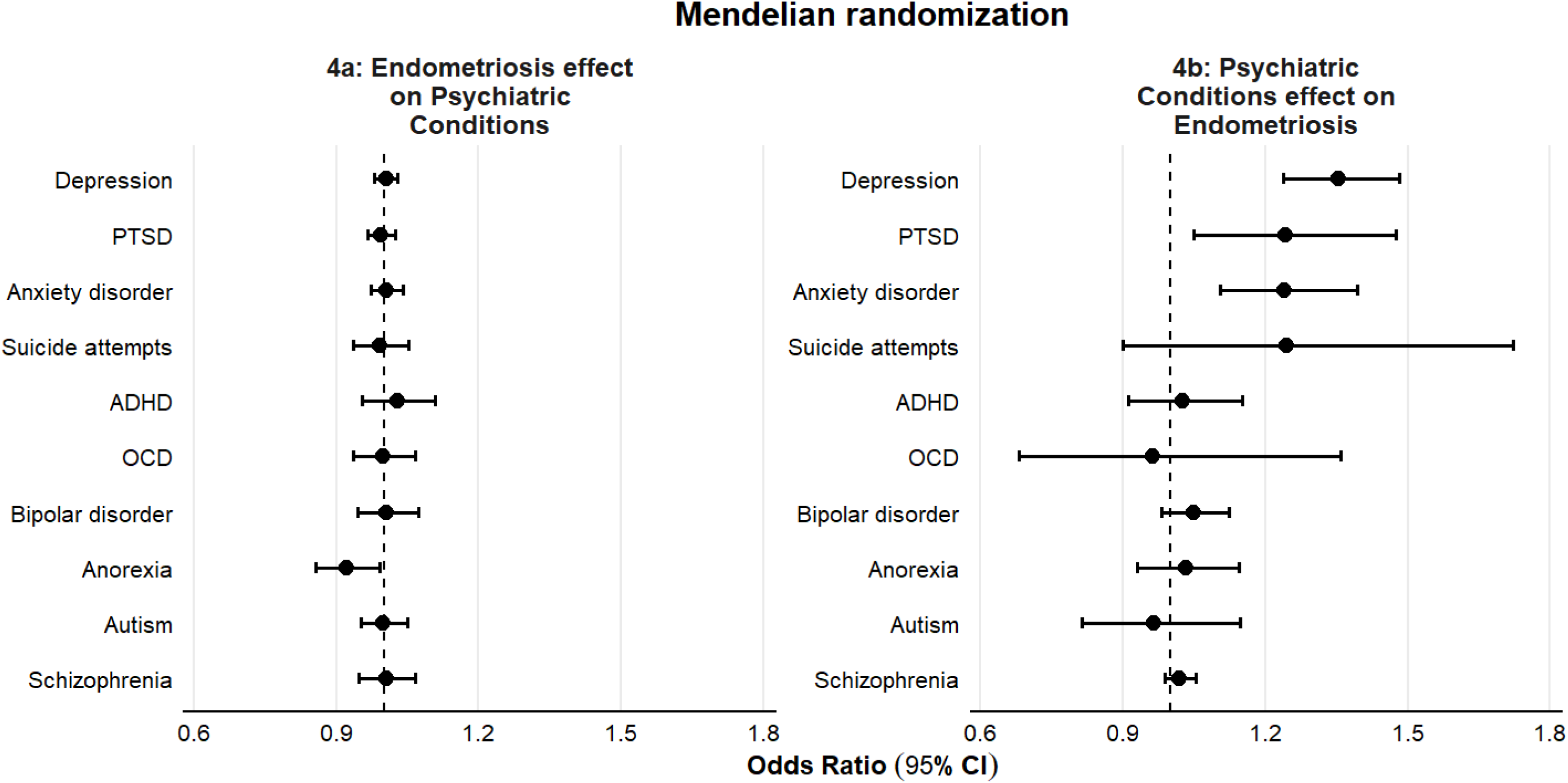
Bidirectional Mendelian randomization of endometriosis and psychiatric conditions. Odds ratios (points) with 95% confidence intervals were derived from the inverse-variance method (or Wald ratio for OCD → endometriosis). The apparent protective effect of endometriosis on anorexia did not survive FDR correction or sensitivity analyses, whereas all other nominal associations remained significant after FDR correction and were consistent across weighted mode, weighted median, and MR-Egger analyses.

**Figure 5.**
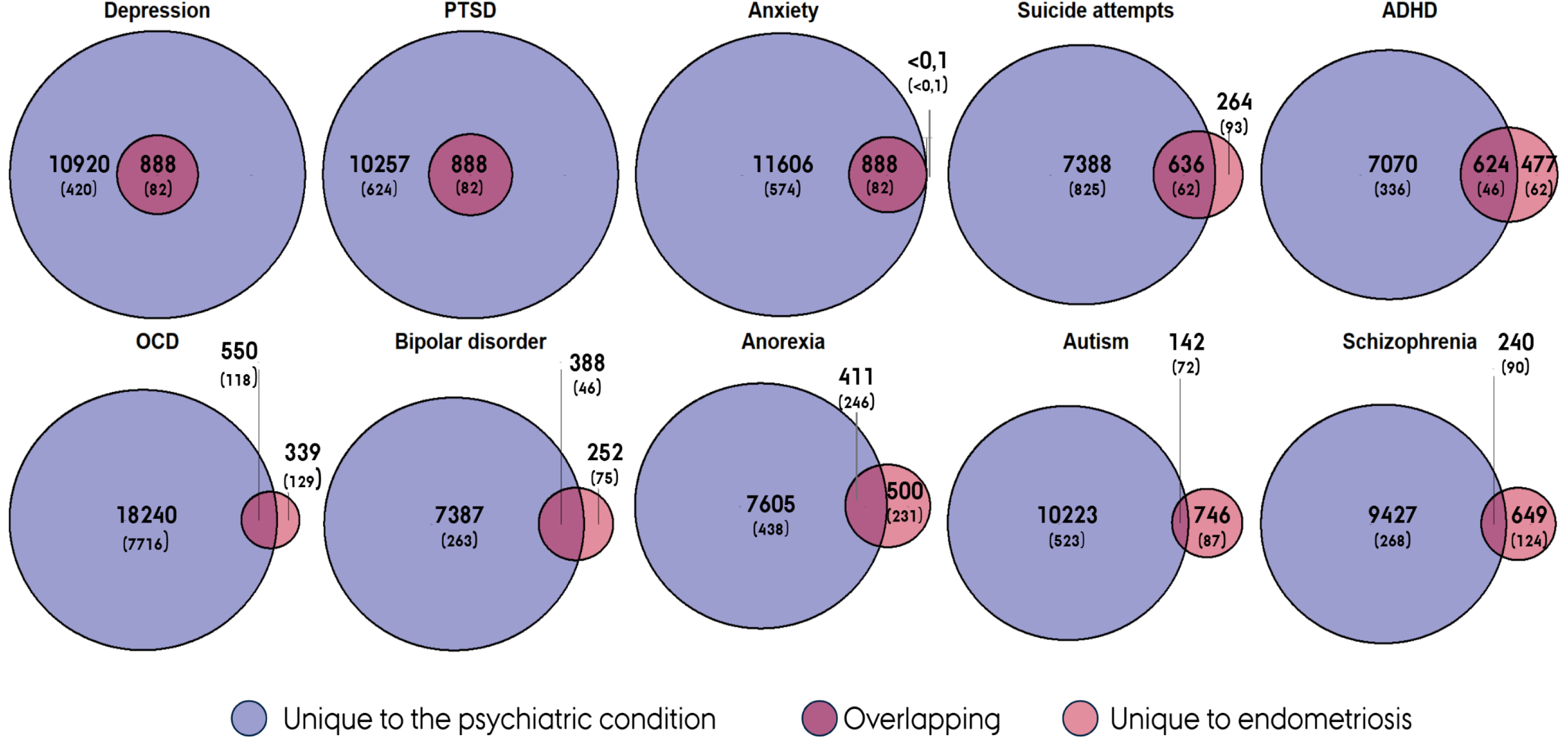
Unique and shared genetic risk markers for endometriosis and psychiatric conditions. Numbers were estimated using bivariate mixture modeling (MiXeR), representing variants that together explain 90% of SNP heritability for each phenotype. Values in brackets indicate the standard deviation of each estimate.

In contrast, we observed consistent evidence that genetic liability to several psychiatric conditions confers an increased risk of endometriosis (**Figure 4b**, Supplementary Table 6). Specifically, genetic liability to depression (OR = 1.36, 95% CI: 1.24–1.50), anxiety (OR = 1.24, 95% CI: 1.11–1.39), and PTSD (OR = 1.30, 95% CI: 1.11–1.53) were each associated with endometriosis risk. All remained statistically significant after FDR-correction. Genetic liability for suicide attempts showed similar point estimates but with wider CIs. These results were consistent across the weighted median, weighted mode, and MR-egger analyses, albeit they often were estimated with greater uncertainty (Supplementary Tables 7 and 8).

There was no evidence of horizontal pleiotropy as indicated by the MR-Egger intercepts (Supplementary Tables 5 and 6). All exposures demonstrated adequate instrument strength, with mean F-statistics exceeding 10 (Supplementary Tables 5 and 6).

### Polygenic overlap - Shared risk markers

To quantify polygenic overlap between endometriosis and psychiatric conditions beyond genetic correlations, we applied bivariate Gaussian mixture modelling (MiXeR) (40). A key observation was the pronounced polygenicity imbalance between endometriosis and psychiatric conditions, with the latter exhibiting approximately 10- to 100-fold greater polygenicity (Supplementary Table 9). Specifically, we estimated that 888 markers explain 90% of the SNP heritability (*h^2^_SNP_*) for endometriosis, compared to between 7,774 and 18,789 markers for psychiatric conditions—the lowest for bipolar disorder and highest for OCD.

Despite this disparity, endometriosis shared a substantial number of risk markers with several psychiatric traits. The strongest overlap was observed for MDD, PTSD, and anxiety; notably, all the 888 risk variants for endometriosis are also found in these conditions, with perfect concordance in effect directions (*ρ*_β_ = >0.999, SD = 3.2× 10⁻⁷ and *ρ*_β_= 1, SD = 8.5 × 10⁻⁷ and *ρ*_β_= 1, SD = 8.0 × 10⁻⁶, respectively). Beyond these, there was high overlap with ADHD and suicide attempts, each with over 600 shared markers and near-perfect concordance in effect direction (*ρ*_β_ = 0.99, SD = 0.017; and *ρ*_β_= 0.98, SD = 0.034, respectively). Bipolar disorder also showed appreciable overlap (388 shared markers; *ρ*_β_ = 0.90, SD = 0.069), while anorexia nervosa and schizophrenia demonstrated more modest sharing, albeit with largely concordant effects. In contrast, autism spectrum disorder exhibited minimal genetic overlap with endometriosis, with only 142 shared markers and markedly discordant effect directions (only 26% concordant). Model fit statistics were consistent with expectation under complete overlap for several conditions (Supplementary Table 9), while for others they were compatible with partial overlap.

### Multivariate GWAS of endometriosis and MDD

Given the strong genetic overlap between endometriosis and mood-related conditions, we focused on MDD and conducted a multivariate GWAS across endometriosis and MDD (**Figure 6a**). Specifically, we applied genomic structural equation modelling (genomic SEM), in which each GWAS was specified to load onto a common latent factor representing shared liability. Overlap between GWAS’s was constrained to operate solely through this factor, ensuring that all shared variance was captured in the common liability. We then estimated each variant’s association with this latent factor. This analysis identified 644 independent genome-wide significant loci (independent at R² < 0,6, distance ≥ 250 kb). Of these, 6% showed significant heterogeneity (Q-heterogeneity P < 0.05), consistent with trait-specific rather than shared effects.

**Figure 6.**
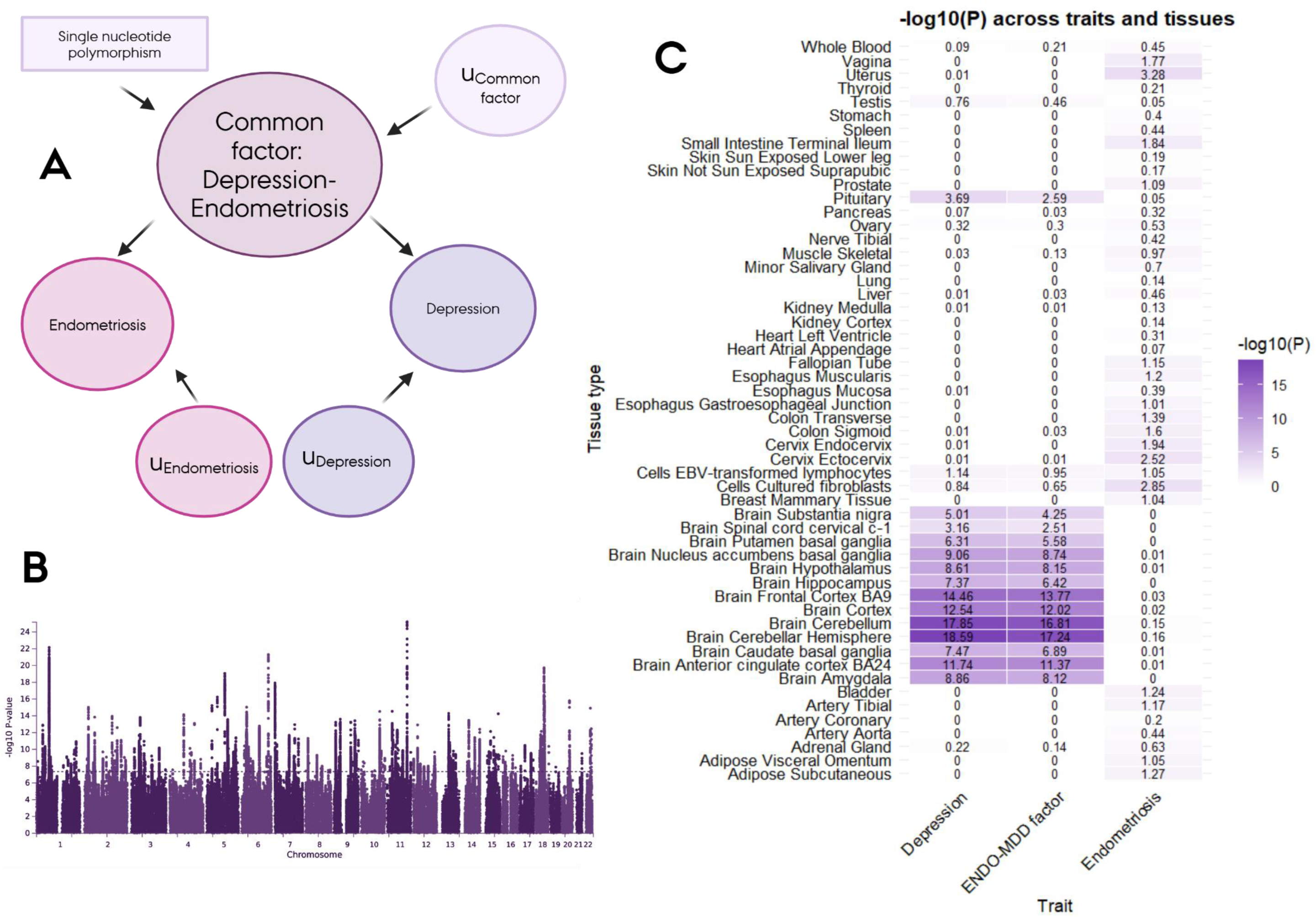
Shared genetic liability between endometriosis and MDD and in which tissues it is expressed. **Figure A** shows the model used to develop the multivariate GWAS using genomic SEM. **Figure B** shows the Manhattan plot of the multivariate GWAS after excluding heterogeneous variants. **Figure C** shows the tissue-specific gene expression of the multivariate GWAS compared with the GWAS for just endometriosis and MDD, examined using MAGMA gene-property analyses across 53 tissue types from GTEx v8.

After excluding heterogeneous variants, 606 independent loci remained GWAS significant as drivers of the common genetic liability between endometriosis and MDD (**Figure 6b**; independent at R² < 0,6, distance ≥ 250 kb; Supplementary Table 10). None of these loci reached genome-wide significance in the original endometriosis GWAS (p-values ranging from 0.04097-0.9968), although all did in the MDD GWAS. The lead association was rs17602038 (P = 6.8×10^−26^), located in *DRD2*, a well-established gene that encodes the D2 subtype of the dopamine receptor, and it has been previously associated with both MDD and schizophrenia and hormone regulation(41–43).

To assess the functional relevance of variants underlying the shared liability between endometriosis and MDD, we examined the non-heterogeneous variants relationship with tissue-specific gene expression (**Figure 6c**). The signals were enriched primarily in brain tissues, closely mirroring patterns observed in the MDD GWAS but less so in the endometriosis GWAS. For example, the strongest signals for both the multivariate GWAS and the MDD GWAS were located in the cerebellum and the cortex, with a similar strength of the associations between the two GWAS. These findings suggest that the shared liability between MDD and endometriosis may be driven largely by convergent brain-related mechanisms commonly seen in MDD.

## Discussion

We find compelling evidence for a shared genetic architecture between endometriosis and several psychiatric conditions, most prominently MDD and related traits. Notably, virtually all endometriosis risk variants were also implicated in MDD, PTSD, and anxiety, and the shared liability appears primarily driven by variants enriched in brain tissues. We observed no evidence that endometriosis causally increases risk for psychiatric conditions. Instead, the opposite was more consistent: genetic liability to psychiatric conditions increased the risk of endometriosis. Together, these findings refine our understanding of the biological links between reproductive and mental health and highlight shared etiological pathways that may represent targets for mechanistic studies and preventive strategies.

### Comparison with previous studies

Consistent with our findings, prior studies have reported global genetic correlations between endometriosis and depression, ranging from r_g_ = 0.27(14) to r_g_ = 0.36(12), as well as correlations observed for anxiety (r_g_ = 0.33(12)), eating disorders (r_g_ = 0.61(12)), and PTSD (r_g_ = 0.31(13)). To our knowledge, no study has previously examined how these correlations vary across the genome. Here, we present evidence which suggest that the genetic overlap is broadly distributed rather than concentrated in a small set of variants: the LAVA analyses point to many weak signals rather than a few strong ones. However, we found a few strongly genetically corelated loci; one in an area associated with somatic mutations common in endometrial cancer and variation in this pathway implicated in schizophrenia, bipolar disorder, and neuroinflammation in major depressive disorder, another in an area with genes involved in estrogen signaling and neurodevelopment, and another locus includes regulators of immune function and hormone-responsive transcription, together highlighting pathways relevant to both endometriosis and psychiatric risk(31–38).

Our MiXeR analyses further suggest that global genetic correlations may underestimate the extent of overlap: because psychiatric conditions are highly polygenic whereas endometriosis is less so, the global rg is attenuated. Strikingly, nearly all variants accounting for the majority of SNP-based heritability (90%) for endometriosis are also represented among the larger set of high-risk variants for mood-related disorders. This pattern is unlikely to reflect polygenicity imbalance alone, as no comparable overlap was observed for autism, which is also highly polygenic but has a distinct etiology from MDD.

The multivariate GWAS of endometriosis and MDD indicated that the shared liability is enriched in brain tissues, with the strongest associations in the cortex and cerebellum, but with significant correlations in all brain related tissues. This parallels findings from analyses of reproductive disorders and MDD in UK Biobank and FinnGen(44). The lead association was located in DRD2, a gene that encodes the D2 subtype of the dopamine receptor, which has been previously associated with both MDD and schizophrenia(41, 42). Taken together, these observations highlight convergent brain-related mechanisms as a plausible basis for the mood-related comorbidities observed in endometriosis.

Previous studies have used MR to probe causal relationships between endometriosis and psychiatric comorbidities. While they examined the effect of psychiatric liability on endometriosis—and, like us, found little evidence of causality—these studies were underpowered to test the clinically presumed reverse direction, namely that endometriosis increases psychiatric risk (e.g., through chronic pain). For example, a recent study estimated the effect at 1.67, but the 95% confidence interval ranged from 0.36 to 2.97, consistent with both risk reduction and risk increase(12). In contrast, our results suggest that genetic liability to endometriosis is unlikely to raise the risk of MDD by more than 3%. This may challenge the prevailing clinical view that psychiatric comorbidity in endometriosis arises primarily as a downstream consequence of the gynecological disorder, especially via pain-related mechanisms.

For the other psychiatric conditions analysed here—suicide attempts, ADHD, OCD, bipolar disorder, autism, and schizophrenia—no prior genetic studies, to our knowledge, have examined their relationships with endometriosis, although epidemiological studies have reported comorbidity with some of these disorders (6). It is plausible that the signals we observe for suicide attempts and ADHD partly reflect their overlap with mood-related traits. However, independent mechanisms unrelated to mood liability cannot be ruled out.

### Considerations

Several considerations should be noted. First, while the recent endometriosis GWAS enabled analyses of this scale, it does not capture the full spectrum of disease(18). Most cases in the GWAS were clinically ascertained, often presenting with severe symptoms such as debilitating pain or endometriosis-related infertility. By contrast, most women with endometriosis remain undiagnosed and never reach the specialist clinics from which such cohorts are drawn. Although the prevalence of endometriosis is estimated at ∼10%, in most populations fewer than 1% of women receive a formal diagnosis(1). While our findings reflect the genetic overlap with psychiatric conditions among clinically ascertained cases, if similar patterns hold for the wider pool of undiagnosed or less severe cases remains to be studied. Expanding reproductive GWAS to more diverse and population-based samples will be essential, and ongoing global initiatives for a reinforced focus on female health research will be critical to this goal(45). Second, although our analysis was more detailed and better powered than prior efforts, it remains restricted to common genetic variants. Rare variants and more complex genetic architectures are increasingly recognized as contributors to both psychiatric and reproductive traits(46, 47). While single rare variants are unlikely to explain population-level risks, they may be enriched in subgroups of women who experience both endometriosis and psychiatric comorbidity, revealing distinct mechanisms and novel intervention targets.

A further limitation is that, to maximize power, we relied on large-scale psychiatric GWAS that were not sex specific(48). A female-specific analysis might reveal more—or less—genetic overlap between endometriosis and psychiatric conditions. Prior efforts restricted to women have not yet been sufficiently powered for analyses of this depth(12), but such work will become increasingly feasible as the scale of biobanks and GWAS expands.

Finally, it is important to emphasize that the observed overlap in genetic risk variants should not be misconstrued as evidence of biological determinism. These findings do not imply that women with endometriosis are destined to develop psychiatric comorbidities. Rather, they offer a framework to understand why comorbidities are common and to identify points for intervention. The shared genetic signals—many enriched in brain tissues—may reflect variation in pain perception, heightened vulnerability to environmental stressors, or other complex mechanisms. Fully disentangling these processes will take time, but such efforts are warranted: clarifying the biological and environmental drivers of psychiatric comorbidity in endometriosis holds real promise for improving diagnosis, treatment, and ultimately the lived experience of affected women.

### Conclusions

We found no evidence that genetic liability to endometriosis increases the risk of psychiatric conditions. In contrast, genetic liability to psychiatric conditions, particularly major depressive disorder and related traits, was associated with increasing the risk of endometriosis. Polygenic analyses revealed extensive shared genetic architecture, with nearly all variants influencing endometriosis also implicated in depression. Leveraging this overlap in a multivariate GWAS, we identify 606 genome-wide significant variants contributing to shared liability and implicate convergent biological pathways—particularly brain-related mechanisms—providing a foundation for mechanistic studies and potential strategies to mitigate psychiatric comorbidity in endometriosis.

## Methods

### Endometriosis and psychiatric genetic data

We used the largest publicly available genome-wide association study (GWAS) summary statistics to conduct genetically informed analyses. GWAS data for endometriosis were obtained from a recent large-scale meta-analysis by Rahmioglu et al(18), which included 60,674 cases and 701,926 controls of European and East Asian ancestry. To minimize population stratification bias, we restricted our analyses to the European subset excluding 23andMe (21,779 cases and 449,087 controls). This study identified 42 genome-wide significant loci encompassing 49 independent association signals, with the strongest signals observed in stage III/IV disease, particularly ovarian endometriosis.

We investigated a broad range of psychiatric conditions previously linked to endometriosis in epidemiological studies(6, 13), using GWAS summary statistics primarily from the Psychiatric Genomics Consortium (PGC). The conditions studied included major depressive disorder (412,305 cases and 1,588,397 controls)(20), PTSD (140,767 cases and 1,109,073 controls)(21), anxiety (120,601 cases and 723,411 controls)(27), suicide attempts (26,590 cases and 492,022 controls)(22), ADHD (38,691 cases and 186,843 controls)(23), OCD (23,493 cases and 1,114,613 controls)(28), bipolar disorder (59,287 cases and 781,022 controls)(24), autism (45,223 cases and 237,910 controls), anorexia (16,992 cases and 55,525 controls)(25) and schizophrenia (67,390 cases and 94,015 controls)(26).

To estimate the specificity of the genetic overlap with depression, we further included GWAS data from a recent meta-analysis of individual depressive symptoms by Adams et al. (19). This study harmonized item-level symptom data from individuals across six cohorts (Psychiatric Genomics Consortium cohorts, Australian Genetics of Depression Study, Generation Scotland, Avon Longitudinal Study of Parents and Children, Estonian Biobank, and UK Biobank) — including both depression case-enriched and community-based samples. The analysis enabled the identification of distinct symptom dimensions and their underlying common variant architecture. Leveraging these data, we examined 11 individual depressive symptoms: hypersomnia, depressed mood, insomnia, fatigue, anhedonia, diminished concentration, recurrent thoughts of death or suicide, weight loss or reduced appetite, feelings of worthlessness or guilt, weight gain or increased appetite, and weight gain in clinical populations. This approach allowed us to assess whether the observed global genetic correlation between endometriosis and depression was concentrated within particular symptom domains.

For details on the studies included and the case ascertainment, see Supplementary Table 1.

### GWAS processing

All GWAS summary statistics were uniformly processed using the *cleansumstats* pipeline (https://github.com/BioPsyk/cleansumstats). For each dataset, we inferred the genome build of the original GWAS and lifted over coordinates to the GRCh37 version of dbSNP. When chromosome and base pair information was unavailable, RSIDs were used instead. The reference allele was aligned to that of the GRCh37 reference genome. Allele directions were flipped as necessary to ensure that the effect allele matched the reference allele, and corresponding effect sizes (e.g., beta coefficients, odds ratios, and z-scores) were adjusted accordingly. Finally, we excluded multi-allelic variants, strand-ambiguous SNPs, allele mismatches, variants with duplicated positions, indels, and those with missing test statistics. The 1000 Genomes Project European reference panel was used for LD estimation.

### Global genetic correlation

We first estimated genome-wide genetic correlations between endometriosis and each psychiatric phenotype using linkage disequilibrium score regression (LDSC)(29). LDSC exploits the relationship between GWAS test statistics and linkage disequilibrium patterns across the genome to estimate genetic correlation (r_g_) with 95% confidence intervals (CI) from summary-level data. Univariate LDSC quality control metrics for our primary phenotypes indicated adequate model fit, with heritability Z-scores >1.5, mean χ² values >1.02, and SNP heritability intercepts between 0.9 and 1.1. We performed false discovery rate (FDR) correction for multiple testing.

### Local genetic correlation

To examine local genetic correlations across the genome, we applied Local Analysis of [co]Variant Association (LAVA) (30). Unlike LDSC regression, which estimates the global genetic correlation across the genome, LAVA partitions the genome into distinct loci and calculates genetic correlations between phenotypes within these regions. This approach addresses the possibility that strong positive correlations in some genomic regions might be masked by negative correlations elsewhere.

A total of 2495 predefined regions across the entire genome were assessed(30). These regions were defined by partitioning the genome into approximately equal-sized blocks (∼1 Mb), while minimizing LD between them(30). Local genetic correlations were tested only in regions showing statistically significant univariate heritability (P < 0.05) for both endometriosis and at least one psychiatric condition. To account for multiple testing, we applied a phenotype-by-phenotype False Discovery Rate (FDR) (q<0.05) correction across the bivariate tested regions.

### Mendelian Randomization

We applied a bidirectional two-sample Mendelian Randomization (2SMR) approach to investigate potential causal associations between endometriosis and psychiatric conditions. This method combines SNP–exposure and SNP–outcome associations within an instrumental variable framework, allowing estimation of directional effects in either direction.(49) Valid instruments must satisfy three core assumptions: (i) strong association with the exposure (relevance), (ii) independence from confounders, and (iii) absence of direct pathways to the outcome other than through the exposure (exclusion restriction).

Instrumental variants were selected based on genome-wide significance (p < 5×10⁻⁸) and LD-clumped (R^2^=0.01 & 10,000 kb window) using a European reference panel to ensure independence. The primary analysis used the inverse-variance weighted (IVW) method; for OCD, where only one independent genome-wide significant SNP was available, we applied the Wald ratio. F-statistics were calculated to assess instrument strength.

All analyses were conducted in R using the *TwoSampleMR* package, the ieugwasr package, the plinkbinr package, and the dplyr package. To complement IVW estimates and account for potential horizontal pleiotropy, we additionally applied the MR-Egger, weighted mode, and weighted median approaches. Horizontal pleiotropy was further assessed via the MR-Egger intercept test.

### Shared risk markers

MiXeR v1.3 is a statistical framework that models the polygenicity and discoverability of complex traits by estimating the number of variants with nonzero additive genetic effects—referred to here as “causal” variants—for each phenotype.(40) Strictly, MiXeR estimates the number of trait-specific and shared variants needed to explain 90% of SNP heritability in each phenotype, irrespective of effect direction.

In accordance with recommendations, we excluded the major histocompatibility complex (chr6:25–34 Mb) from all analyses. For each trait and trait pair, we randomly sampled two million SNPs with minor allele frequency ≥5% and repeated model fitting 20 times to assess convergence and quantify the variability of parameter estimates. This procedure follows established recommendations to improve model stability and reproducibility.

To assess model fit, we compared the Akaike Information Criterion across nested models. For univariate analyses, we compared the MiXeR model to a baseline LD score regression–based model lacking a polygenicity parameter. For bivariate analyses, we evaluated fit across three models: no overlap, partial overlap (as estimated), and full overlap (all causal variants of the less polygenic trait are shared with the other). Model diagnostics and consistency across replicates were used to assess performance (Supplementary Table 7).

### Multivariate GWAS and tissue-enrichment

Given the high genetic overlap between endometriosis and mood-related conditions, we focused on MDD and endometriosis to characterize shared liability. We applied genomic structural equation modeling (GSEM) to perform a multivariate GWAS(50), fitting a model in which both traits load onto a single common factor, with each SNP regressed onto this factor and residual variance assumed independent of the shared liability. This approach provides estimates of each SNP’s contribution to the unit-scaled shared genetic liability. LDSC performed on the resulting summary statistics indicated minimal genomic inflation (LDSC quality control metrics indicated adequate model fit, with heritability Z-score >1.5, mean χ² value >1.02, and SNP heritability intercept between 0.9 and 1.1). Independent loci were defined using PLINK v1.9 within FUMA, with R² ≥ 0.6 and distance ≥ 250 kb. To isolate variants consistent with the shared factor rather than trait-specific effects, SNPs with a Q-heterogeneity p-value < 0.05 were excluded, leaving variants that reflect shared liability between endometriosis and MDD (post-filter LDSC quality control metrics indicated adequate model fit, with heritability Z-score 31, mean χ² value 2.07, and SNP heritability intercept 1.02).

We next used FUMA to investigate tissue specificity of the associated variants. Tissue enrichment was assessed with MAGMA gene-property analyses(51), which test the relationship between tissue-specific gene expression profiles and disease–gene associations. Analyses were performed across 53 tissue types from GTEx v8(52). To compare tissue expression patterns of the shared liability with those of MDD and endometriosis individually, we repeated the same analyses using the original GWAS data.

## Data Availability

All data produced in the present study are available upon reasonable request to the authors and after necessary approvals from PGC.

