## Supplementary Table 4 for "Shared Genetic Architecture Between Endometriosis and Psychiatric Conditions May Explain Comorbidity"

### STable 4 Local genetic correlation between endometriosis and psychiatric conditions using LAVA

| **Psychiatric condition** | **Number of bivariate correlated loci** | **Number of positively correlated loci** | **Number of negatively correlated loci** | **Number of positive statistically significant (p<0.05, unadjusted) correlated loci** | **Number of negative statistically significant (p<0.05, unadjusted) correlated loci** | **Number of FDR statistically significantly (q<0.05) correlated loci** | **Number of positive FDR statistically significantly (q<0.05) correlated loci** | **Number of negative FDR statistically significantly (q<0.05) correlated loci** |
| --- | --- | --- | --- | --- | --- | --- | --- | --- |
| Depression | 202 | 135 | 67 | 35 | 5 | 0 | 0 | 0 |
| PTSD | 123 | 81 | 42 | 10 | 7 | 0 | 0 | 0 |
| Anxiety disorder | 145 | 86 | 59 | 12 | 5 | 0 | 0 | 0 |
| Suicide attempts | 110 | 59 | 51 | 6 | 4 | 2 | 2 | 0 |
| ADHD | 118 | 67 | 51 | 5 | 3 | 0 | 0 | 0 |
| OCD | 88 | 60 | 28 | 6 | 6 | 1 | 1 | 0 |
| Bipolar disorder | 151 | 98 | 53 | 12 | 5 | 0 | 0 | 0 |
| Anorexia | 84 | 45 | 39 | 6 | 5 | 0 | 0 | 0 |
| Autism | 88 | 43 | 45 | 2 | 3 | 0 | 0 | 0 |
| Schizophrenia | 167 | 88 | 79 | 15 | 5 | 3 | 1 | 2 |
