## Supplementary Table 5 for "Shared Genetic Architecture Between Endometriosis and Psychiatric Conditions May Explain Comorbidity"

### STable 5 MR-analysis of endometriosis on psychiatric conditions using Inverse-variance weighted

| **Exposure** | **Outcome** | **Number of SNPs** | **Odds Ratio** | **95% CI** | **P-value** | **FDR-corrected**  **p** | **Egger intercept** | **Egger se** | **Egger pval** | **Mean F** |
| --- | --- | --- | --- | --- | --- | --- | --- | --- | --- | --- |
| endometriosis | Depression | 22 | 1,005 | 0.981 to 1.029 | 0,699 | 0.983 | 0,004 | 0,004 | 0,243 | 47,85 |
|  | PTSD | 20 | 0,994 | 0.965 to 1.025 | 0,718 | 0.983 | 0,003 | 0,005 | 0,541 | 49,33 |
|  | Anxiety disorder | 23 | 1,006 | 0.974 to 1.04 | 0,695 | 0.983 | -0,002 | 0,005 | 0,692 | 47,16 |
|  | Suicide attempts | 20 | 0,992 | 0.935 to 1.053 | 0,791 | 0.983 | -0,001 | 0,01 | 0,893 | 45,85 |
|  | ADHD | 21 | 1,029 | 0.955 to 1.108 | 0,457 | 0.983 | 0,006 | 0,012 | 0,6 | 48,55 |
|  | OCD | 23 | 0,999 | 0.936 to 1.065 | 0,969 | 0.983 | -0,0007 | 0,01 | 0,940 | 47,16 |
|  | Bipolar disorder | 20 | 1,006 | 0.944 to 1.073 | 0,846 | 0.983 | -0,020 | 0,009 | 0,046 | 49,37 |
|  | Anorexia | 21 | 0,921 | 0.856 to 0.992 | 0,029 | 0.289 | 0,012 | 0,011 | 0,303 | 48,55 |
|  | Autism | 23 | 0,999 | 0.951 to 1.05 | 0,983 | 0.983 | -0,010 | 0,007 | 0,210 | 47,16 |
|  | Schizophrenia | 23 | 1,005 | 0.948 to 1.066 | 0,856 | 0.983 | 0,008 | 0,009 | 0,366 | 47,16 |
