## Supplementary Table 6 for "Shared Genetic Architecture Between Endometriosis and Psychiatric Conditions May Explain Comorbidity"

### STable 6 MR-analysis of psychiatric conditions on endometriosis risks using Inverse-variance weighted or Wald ratio

| **Exposure** | **Outcome** | **Number of SNPs** | **Odds Ratio** | **95% CI** | **P-value** | **FDR-corrected**  **p** | **Egger intercept** | **Egger se** | **Egger pval** | **Mean F** |
| --- | --- | --- | --- | --- | --- | --- | --- | --- | --- | --- |
| Depression | endometriosis | 252 | 1,355 | 1.238 to 1.482 | 3.55 × 10⁻¹¹ | 3,546× 10⁻^10^ | 0,006 | 0,004 | 0,188 | 42,67 |
| PTSD | endometriosis | 27 | 1,243 | 1.048 to 1.475 | 0.013 | 0,042 | 0,017 | 0,011 | 0,135 | 39,32 |
| Anxiety disorder | endometriosis | 58 | 1,24 | 1.105 to 1.393 | 2.68 × 10⁻⁴ | 0,001 | -0,003 | 0,008 | 0,755 | 36,8 |
| Suicide attempts | endometriosis | 2 | 1,245 | 0.899 to 1.723 | 0,187 | 0,374 | - | - | - | 37,57 |
| ADHD | endometriosis | 25 | 1,025 | 0.912 to 1.152 | 0,679 | 0,764 | 0,009 | 0,019 | 0,635 | 38,8 |
| OCD* | endometriosis | 1 | 0,962 | 0.682 to 1.358 | 0,826 | 0,826 | - | - | - | 38,2 |
| Bipolar disorder | endometriosis | 75 | 1,05 | 0.981 to 1.123 | 0,157 | 0,374 | -0,004 | 0,009 | 0,682 | 37,66 |
| Anorexia | endometriosis | 7 | 1,033 | 0.931 to 1.145 | 0,544 | 0,764 | 0,0129 | 0,020 | 0,539 | 36,9 |
| Autism | endometriosis | 11 | 0,965 | 0.813 to 1.146 | 0,688 | 0,764 | -0,028 | 0,026 | 0,303 | 40,28 |
| Schizophrenia | endometriosis | 175 | 1,02 | 0.988 to 1.054 | 0,230 | 0,383 | 0,004 | 0,004 | 0,426 | 43,82 |

*Wald ratio
