## Supplementary Table 7 for "Shared Genetic Architecture Between Endometriosis and Psychiatric Conditions May Explain Comorbidity"

### STable 7 MR-analysis of psychiatric conditions on endometriosis sensitivity analyses

| **Exposure** | **Method** | **Number of SNPs** | **Odds Ratio** | **95% CI** | **P-value** |
| --- | --- | --- | --- | --- | --- |
| Depression | MR Egger | 252 | 1,038 | 0.693 to 1.557 | 0.855 |
| Depression | Weighted median | 252 | 1,366 | 1.22 to 1.529 | 6.06 × 10⁻⁸ |
| Depression | Inverse variance weighted | 252 | 1,355 | 1.238 to 1.482 | 3.55 × 10⁻¹¹ |
| Depression | Simple mode | 252 | 1,307 | 0.88 to 1.942 | 0.186 |
| Depression | Weighted mode | 252 | 1,356 | 0.943 to 1.951 | 0.102 |
| PTSD | MR Egger | 27 | 0,718 | 0.351 to 1.469 | 0.373 |
| PTSD | Weighted median | 27 | 1,252 | 0.994 to 1.577 | 0.056 |
| PTSD | Inverse variance weighted | 27 | 1,243 | 1.048 to 1.475 | 0.013 |
| PTSD | Simple mode | 27 | 1,671 | 1.052 to 2.653 | 0.039 |
| PTSD | Weighted mode | 27 | 1,475 | 0.934 to 2.328 | 0.107 |
| Anxiety | MR Egger | 58 | 1,334 | 0.834 to 2.133 | 0.234 |
| Anxiety | Weighted median | 58 | 1,228 | 1.064 to 1.419 | 0.005 |
| Anxiety | Inverse variance weighted | 58 | 1,24 | 1.105 to 1.393 | 0.000 |
| Anxiety | Simple mode | 58 | 1,271 | 0.904 to 1.785 | 0.173 |
| Anxiety | Weighted mode | 58 | 1,254 | 0.899 to 1.748 | 0.187 |
| Suicide attempts | Inverse variance weighted | 2 | 1,245 | 0.899 to 1.723 | 0.187 |
| ADHD | MR Egger | 25 | 0,898 | 0.517 to 1.559 | 0.706 |
| ADHD | Weighted median | 25 | 1,123 | 0.994 to 1.27 | 0.063 |
| ADHD | Inverse variance weighted | 25 | 1,025 | 0.912 to 1.152 | 0.679 |
| ADHD | Simple mode | 25 | 1,173 | 0.948 to 1.453 | 0.155 |
| ADHD | Weighted mode | 25 | 1,173 | 0.921 to 1.494 | 0.207 |
| OCD | Wald ratio | 1 | 0,962 | 0.682 to 1.358 | 0.826 |
| Bipolar disorder | MR Egger | 75 | 1,128 | 0.796 to 1.599 | 0.501 |
| Bipolar disorder | Weighted median | 75 | 1,065 | 0.983 to 1.153 | 0.121 |
| Bipolar disorder | Inverse variance weighted | 75 | 1,05 | 0.981 to 1.123 | 0.157 |
| Bipolar disorder | Simple mode | 75 | 1,108 | 0.893 to 1.373 | 0.355 |
| Bipolar disorder | Weighted mode | 75 | 1,089 | 0.896 to 1.322 | 0.394 |
| Autism | MR Egger | 11 | 1,471 | 0.677 to 3.195 | 0.355 |
| Autism | Weighted median | 11 | 1,018 | 0.865 to 1.197 | 0.833 |
| Autism | Inverse variance weighted | 11 | 0,965 | 0.813 to 1.146 | 0.688 |
| Autism | Simple mode | 11 | 1,051 | 0.797 to 1.386 | 0.732 |
| Autism | Weighted mode | 11 | 1,036 | 0.83 to 1.293 | 0.763 |
| Anorexia | MR Egger | 7 | 0,904 | 0.601 to 1.36 | 0.648 |
| Anorexia | Weighted median | 7 | 1,037 | 0.906 to 1.188 | 0.599 |
| Anorexia | Inverse variance weighted | 7 | 1,033 | 0.931 to 1.145 | 0.544 |
| Anorexia | Simple mode | 7 | 1,116 | 0.905 to 1.376 | 0.343 |
| Anorexia | Weighted mode | 7 | 1,079 | 0.879 to 1.323 | 0.495 |
| Schizophrenia | MR Egger | 175 | 0,967 | 0.846 to 1.106 | 0.629 |
| Schizophrenia | Weighted median | 175 | 1,027 | 0.982 to 1.075 | 0.245 |
| Schizophrenia | Inverse variance weighted | 175 | 1,02 | 0.988 to 1.054 | 0.230 |
| Schizophrenia | Simple mode | 175 | 1,063 | 0.931 to 1.214 | 0.368 |
| Schizophrenia | Weighted mode | 175 | 1,038 | 0.917 to 1.176 | 0.554 |
