## Supplementary Table 8 for "Shared Genetic Architecture Between Endometriosis and Psychiatric Conditions May Explain Comorbidity"

### STable 8 MR-analysis of endometriosis on psychiatric conditions sensitivity analyses

| **Outcome** | **Method** | **Number of SNPs** | **Odds Ratio** | **95% CI** | **P-value** |
| --- | --- | --- | --- | --- | --- |
| Depression | MR Egger | 22 | 0,962 | 0.893 to 1.036 | 0.320 |
| Depression | Weighted median | 22 | 1,004 | 0.981 to 1.028 | 0.714 |
| Depression | Inverse variance weighted | 22 | 1,005 | 0.981 to 1.029 | 0.699 |
| Depression | Simple mode | 22 | 1,01 | 0.971 to 1.051 | 0.619 |
| Depression | Weighted mode | 22 | 1,004 | 0.974 to 1.035 | 0.796 |
| PTSD | MR Egger | 20 | 0,965 | 0.874 to 1.066 | 0.491 |
| PTSD | Weighted median | 20 | 0,995 | 0.96 to 1.032 | 0.791 |
| PTSD | Inverse variance weighted | 20 | 0,994 | 0.965 to 1.025 | 0.718 |
| PTSD | Simple mode | 20 | 1,011 | 0.95 to 1.075 | 0.742 |
| PTSD | Weighted mode | 20 | 1,004 | 0.943 to 1.069 | 0.898 |
| Anxiety | MR Egger | 23 | 1,028 | 0.924 to 1.143 | 0.621 |
| Anxiety | Weighted median | 23 | 0,989 | 0.95 to 1.029 | 0.579 |
| Anxiety | Inverse variance weighted | 23 | 1,006 | 0.974 to 1.04 | 0.695 |
| Anxiety | Simple mode | 23 | 0,981 | 0.9 to 1.069 | 0.660 |
| Anxiety | Weighted mode | 23 | 0,98 | 0.922 to 1.041 | 0.513 |
| Suicide attempts | MR Egger | 20 | 1,007 | 0.808 to 1.254 | 0.953 |
| Suicide attempts | Weighted median | 20 | 0,963 | 0.885 to 1.048 | 0.378 |
| Suicide attempts | Inverse variance weighted | 20 | 0,992 | 0.935 to 1.053 | 0.791 |
| Suicide attempts | Simple mode | 20 | 0,954 | 0.814 to 1.119 | 0.570 |
| Suicide attempts | Weighted mode | 20 | 0,943 | 0.835 to 1.065 | 0.356 |
| ADHD | MR Egger | 21 | 0,963 | 0.747 to 1.241 | 0.774 |
| ADHD | Weighted median | 21 | 1,018 | 0.937 to 1.106 | 0.668 |
| ADHD | Inverse variance weighted | 21 | 1,029 | 0.955 to 1.108 | 0.457 |
| ADHD | Simple mode | 21 | 0,988 | 0.837 to 1.166 | 0.886 |
| ADHD | Weighted mode | 21 | 0,978 | 0.842 to 1.136 | 0.775 |
| OCD | MR Egger | 23 | 1,006 | 0.816 to 1.242 | 0.952 |
| OCD | Weighted median | 23 | 0,989 | 0.904 to 1.081 | 0.807 |
| OCD | Inverse variance weighted | 23 | 0,999 | 0.936 to 1.065 | 0.969 |
| OCD | Simple mode | 23 | 0,947 | 0.807 to 1.112 | 0.514 |
| OCD | Weighted mode | 23 | 0,937 | 0.8 to 1.097 | 0.425 |
| Bipolar disorder | MR Egger | 20 | 1,236 | 1.015 to 1.504 | 0.049 |
| Bipolar disorder | Weighted median | 20 | 1,009 | 0.945 to 1.077 | 0.795 |
| Bipolar disorder | Inverse variance weighted | 20 | 1,006 | 0.944 to 1.073 | 0.846 |
| Bipolar disorder | Simple mode | 20 | 0,993 | 0.873 to 1.13 | 0.921 |
| Bipolar disorder | Weighted mode | 20 | 1,009 | 0.903 to 1.128 | 0.874 |
| Autism | MR Egger | 23 | 1,103 | 0.942 to 1.291 | 0.236 |
| Autism | Weighted median | 23 | 0,997 | 0.931 to 1.068 | 0.943 |
| Autism | Inverse variance weighted | 23 | 0,999 | 0.951 to 1.05 | 0.983 |
| Autism | Simple mode | 23 | 0,962 | 0.845 to 1.096 | 0.567 |
| Autism | Weighted mode | 23 | 0,962 | 0.863 to 1.073 | 0.496 |
| Anorexia | MR Egger | 21 | 0,812 | 0.635 to 1.038 | 0.113 |
| Anorexia | Weighted median | 21 | 0,917 | 0.828 to 1.016 | 0.098 |
| Anorexia | Inverse variance weighted | 21 | 0,921 | 0.856 to 0.992 | 0.029 |
| Anorexia | Simple mode | 21 | 0,889 | 0.74 to 1.068 | 0.224 |
| Anorexia | Weighted mode | 21 | 0,906 | 0.775 to 1.06 | 0.232 |
| Schizophrenia | MR Egger | 23 | 0,924 | 0.764 to 1.116 | 0.421 |
| Schizophrenia | Weighted median | 23 | 1,042 | 0.973 to 1.115 | 0.237 |
| Schizophrenia | Inverse variance weighted | 23 | 1,005 | 0.948 to 1.066 | 0.856 |
| Schizophrenia | Simple mode | 23 | 1,065 | 0.923 to 1.229 | 0.400 |
| Schizophrenia | Weighted mode | 23 | 1,065 | 0.935 to 1.213 | 0.355 |
