## Supplementary Table 9 for "Shared Genetic Architecture Between Endometriosis and Psychiatric Conditions May Explain Comorbidity"

### STable 9 MiXeR results

| Trait 1 | Endometriosis | Endometriosis | Endometriosis | Endometriosis | Endometriosis | Endometriosis | Endometriosis | Endometriosis | Endometriosis | Endometriosis |
| --- | --- | --- | --- | --- | --- | --- | --- | --- | --- | --- |
| Trait 2 | Depression | PTSD | Anxiety | Suicide attempts | ADHD | OCD | Anorexia | Bipolar disorder | Autism | Schizophrenia |
| Dice coefficient (mean) | 0.140 | 0.148 | 0.133 | 0.143 | 0.145 | 0.058 | 0.091 | 0.09 | 0.025 | 0.046 |
| Dice coefficient (standard deviation) | 0.013 | 0.014 | 0.014 | 0.014 | 0.011 | 0.016 | 0.052 | 0.012 | 0.012 | 0.017 |
| Polygenicity of Trait 1 (π₁ mean) | 1.276e-10 | 1.585e-10 | 3.869e-10 | 7.91E-05 | 8.30E-05 | 1.062e-04 | 1.50E-04 | 1.57E-04 | 2.34E-04 | 2.03E-04 |
| Polygenicity of Trait 1 (π₁ SD) | 1.515e-10 | 1.682e-10 | 6.466e-10 | 2.90E-05 | 1.93E-05 | 4.055e-05 | 7.26E-05 | 2.35E-05 | 2.72E-05 | 3.89E-05 |
| Polygenicity of Trait 2 (π₂ mean) | 0.003 | 0.003 | 0.004 | 0.002 | 0.002 | 0.006 | 0.002 | 0.002 | 0.003 | 0.003 |
| Polygenicity of Trait 2 (π₂ SD) | 1.284e-04 | 1.956e-04 | 1.799e-04 | 2.59E-04 | 1.05E-04 | 0.001 | 1.37E-04 | 8.24E-05 | 1.64E-04 | 8.41E-05 |
| Shared polygenicity (π₁₂ mean) | 2.785e-04 | 2.785e-04 | 2.785e-04 | 1.99E-04 | 1.96E-04 | 1.723e-04 | 1.29E-04 | 1.22E-04 | 4.45E-05 | 7.52E-05 |
| Shared polygenicity (π₁₂ SD) | 2.571e-05 | 2.571e-05 | 2.571e-05 | 1.95E-05 | 1.44E-05 | 3.698e-05 | 7.70E-05 | 1.45E-05 | 2.25E-05 | 2.82E-05 |
| Number of causal variants in Trait 1 at p < 1e-9 (mean) | 4.07e-04 | 5.06e-04 | 0.001 | 252.3 | 264.7 | 338.716 | 476.9 | 500.0 | 746.3 | 648.5 |
| Number of causal variants in Trait 1 at p < 1e-9 (SD) | 4.83e-04 | 5.364e-04 | 0.002 | 92.5 | 61.7 | 129.332 | 231.4 | 74.9 | 86.8 | 124.0 |
| Number of causal variants in Trait 2 at p < 1e-9 (mean) | 10919.8 | 10257.2 | 11606.5 | 7387.6 | 7070.3 | 18239.823 | 7604.8 | 7386.5 | 10223.0 | 9427.1 |
| Number of causal variants in Trait 2 at p < 1e-9 (SD) | 409.5 | 623.9 | 573.6 | 825.4 | 336.2 | 4716.199 | 437.8 | 262.8 | 522.6 | 268.1 |
| Number of shared causal variants at p < 1e-9 (mean) | 888.3 | 888.3 | 888.3 | 636.0 | 623.6 | 549.594 | 411.4 | 388.3 | 142.0 | 239.8 |
| Number of shared causal variants at p < 1e-9 (SD) | 82.0 | 82.0 | 82.0 | 62.1 | 46.0 | 117.921 | 245.5 | 46.3 | 71.7 | 89.8 |
| Zero-effect correlation (ρ₀ mean) | 0.039 | 0.031 | 0.027 | 0.024 | 0.019 | 0.007 | 0.006 | 0.025 | 0.004 | 0.024 |
| Zero-effect correlation (ρ₀ SD) | 0.002 | 0.002 | 0.002 | 0.002 | 0.002 | 0.002 | 0.002 | 0.002 | 0.002 | 0.002 |
| Effect size correlation among shared variants (ρ_β mean) | >0.999 | 1.000 | 1.0 | 0.977 | 0.986 | 0.839 | 0.539 | 0.902 | -0.643 | 0.593 |
| Effect size correlation among shared variants (ρ_β SD) | 3.205e-07 | 8.453e-07 | 8.007e-06 | 0.034 | 0.017 | 0.138 | 0.332 | 0.069 | 0.254 | 0.19 |
| Genetic correlation (r₉ mean) | 0.274 | 0.282 | 0.267 | 0.233 | 0.235 | 0.112 | 0.058 | 0.133 | -0.028 | 0.045 |
| Genetic correlation (r₉ SD) | 0.013 | 0.015 | 0.015 | 0.018 | 0.012 | 0.019 | 0.019 | 0.011 | 0.014 | 0.013 |
| Fraction of concordant effect directions among shared variants (mean) | >0.999 | 1.000 | 0.999 | 0.943 | 0.953 | 0.839 | 0.709 | 0.867 | 0.261 | 0.71 |
| Fraction of concordant effect directions among shared variants (SD) | 1.351e-04 | 2.077e-04 | 0.001 | 0.037 | 0.027 | 0.09 | 0.151 | 0.051 | 0.11 | 0.08 |
| ΔAIC (best vs. minimum AIC across models) | -1.993 | -1.992 | -1.967 | -0.47 | -0.73 | -0.531 | -0.56 | -0.34 | -0.34 | 0.98 |
| ΔBIC (best vs. minimum BIC across models) | -11.498 | -11.459 | -11.512 | -10.00 | -10.11 | -9.963 | -9.93 | -9.90 | -9.90 | -8.56 |
| ΔAIC (best vs. maximum AIC across models) | -1.967 | -1.985 | -1.984 | 4.36 | 6.79 | 0.226 | 0.06 | 5.44 | 4.85 | 8.95 |
| ΔBIC (best vs. maximum BIC across models) | -11.472 | -11.452 | -11.529 | -5.17 | -2.59 | -9.205 | -9.31 | -4.11 | -4.70 | -0.58 |
