## Supplementary Table 10 for "Shared Genetic Architecture Between Endometriosis and Psychiatric Conditions May Explain Comorbidity"

### STable 10 FUMA results

| #Genomic risk loci | 193 |
| --- | --- |
| #lead SNPs | 248 |
| #Ind. Sig. SNPs | 606 |
| #candidate SNPs | 35482 |
| #candidate GWAS tagged SNPs | 19530 |
| #mapped genes | 527 |
