## Supplementary Table 1 for "Shared Genetic Architecture Between Endometriosis and Psychiatric Conditions May Explain Comorbidity"

### STable 1 Sources of GWAS data

| **Phenotype** | **Study/Consortium** | **Phenotype assessment** | **Year** | **N cases** | **N controls** |
| --- | --- | --- | --- | --- | --- |
| Endometriosis | Rahmioglu et al | Mix of surgically confirmed, medical records, and self-reported | 2023 | 21 779 | 449 087 |
| Depression | Adams et al, PCG | Mix of structured diagnostic interviews and self-reports of MDD diagnosis | 2025 | 412 305 | 1 588 397 |
| Depression symptoms | Adams et al | Structured diagnostic interviews or clinical assessments of their current or lifetime worst episode. Or history of receiving treatment for depression and assessed symptoms during their worst episode using an online questionnaire. These were grouped together as ‘Case-enriched’ cohorts. Current depressive symptoms collected by interview, depression symptoms from worst episode using online surveys. Symptom data in these two cohorts had a skip-structure, where all participants were asked about mood and anhedonia symptoms while only participants who endorsed at least one cardinal symptom were asked about the other DSM symptoms. Retrospective assessments of prolonged low mood and/or anhedonia. These were included regardless of depression diagnosis and were grouped together as ‘Community cohorts’. | 2024 | - | - |
| PTSD | Nievergelt et al | Mix of interviews types and scales | 2024 | 140767 | 1109073 |
| Anxiety disorder | Strom et al | The majority of the sample (70%) had completed the GAD-7 or closely related brief self-report measures assessing recent anxiety symptoms. The remaining 30% used other brief self-report anxiety scales. | 2024 | 120601 | 723411 |
| Suicide attempts | Mullins et al | Cases were individuals who made a nonfatal SA or died by suicide. A nonfatal SA was defined as a lifetime act of deliberate self-harm with the intent to die. Information on SA was ascertained using structured clinical interviews, self-report questionnaires, and hospital records or International Classification of Diseases codes. A proportion of cases in the iPSYCH and Columbia University cohorts had died by suicide, determined using the Cause of Death Register in Denmark and the Columbia Classification Algorithm for Suicide Assessment, respectively | 2022 | 26 590 | 492 022 |
| ADHD | Demontis et al | ICD diagnoses, prescription of ADHD medication | 2023 | 38 691 | 186 843 |
| OCD | Strom et al | Some cases met DSM-5 or ICD-10 criteria for OCD as assessed by a healthcare professional or derived from (electronic) health records, while the remaining cases were based on self-reported OCD diagnosis. | 2025 | 23 493 | 1 114 613 |
| Bipolar disorder | O'Connell et al | Mix of semi-structured clinical interviews, medical records, registries, questionnaire data, and self-reported surveys | 2025 | 59 287 | 78 1022 |
| Anorexia nervosa | Watson et al | Lifetime diagnosis of anorexia nervosa via hospital or register records, structured clinical interviews, self-reported anorexia diagnosis, or online questionnaires based on standardized criteria | 2019 | 16 992 | 55 525 |
| Autism | Autism Spectrum Disorder Working Group of the Psychiatric Genomics Consortium | Clinically Diagnosed | 2023 | 45223 | 237910 |
| Schizophrenia | Trubetskoy et al | Consensus diagnosis, research diagnostic interview, review of medical records, mixed  strategy | 2022 | 67 390 | 94 015 |
