## Supplementary Table 2 for "Shared Genetic Architecture Between Endometriosis and Psychiatric Conditions May Explain Comorbidity"

### STable 2 Genetic correlation between endometriosis and different psychiatric conditions using LDSC

| **Psychiatric condition** | **rg** | **se** | **z** | **p** | **FDR-corrected**  **p** |
| --- | --- | --- | --- | --- | --- |
| Depression | 0.357 | 0.031 | 11.475 | 1.75 × 10⁻³⁰ | 1.75 × 10⁻²⁹ |
| PTSD | 0.335 | 0.040 | 8.384 | 5.12 × 10⁻¹⁷ | 2.56 × 10⁻¹⁶ |
| Anxiety disorder | 0.290 | 0.036 | 8.095 | 5.73 × 10⁻¹⁶ | 1.91 × 10⁻¹⁵ |
| Suicide attempts | 0.260 | 0.061 | 4.275 | 1.91 × 10⁻⁵ | 3.83 × 10⁻⁵ |
| ADHD | 0.206 | 0.046 | 4.463 | 8.09 × 10⁻⁶ | 2.02 × 10⁻⁵ |
| OCD | 0.131 | 0.053 | 2.489 | 1.28 × 10⁻² | 0.018 |
| Bipolar disorder | 0.131 | 0.041 | 3.212 | 1.30 × 10⁻³ | 0.002 |
| Anorexia nervosa | 0.056 | 0.046 | 1.230 | 0.219 | 0.243 |
| Autism | 0.043 | 0.054 | 0.796 | 0.426 | 0,473 |
| Schizophrenia | 0.018 | 0.033 | 0.559 | 0.577 | 0.577 |
