## Supplementary Table 3 for "Shared Genetic Architecture Between Endometriosis and Psychiatric Conditions May Explain Comorbidity"

### STable 3 Genetic correlation between endometriosis and depression symptoms using LDSC

| **Depression symptoms**  community **=** symptoms, not necessarily clinical MDD diagnosis  clinical = diagnosis of MDD | **cases** | **controls** | **rg** | **se** | **p** | **FDR-corrected**  **p** |
| --- | --- | --- | --- | --- | --- | --- |
| Increased sleep | 20125 | 20055 | 0.275 | 0.090 | 2.00 × 10⁻³ | 0,008 |
| Depressed mood | 107956 | 99480 | 0.254 | 0.044 | 1.12 × 10⁻⁸ | 1,23× 10⁻^7^ |
| Sleep problems | 73144 | 19851 | 0.231 | 0.090 | 0.010 | 0,023 |
| Fatigue | 85304 | 16736 | 0.231 | 0.085 | 0.007 | 0,018 |
| Anhedonia | 81113 | 126167 | 0.202 | 0.044 | 4.70 × 10⁻⁶ | 2,584× 10⁻^5^ |
| Trouble concentrating | 75190 | 23416 | 0.1960 | 0.094 | 0.038 | 0,052 |
| Thoughts of wanting to die | 46984 | 58885 | 0.191 | 0.092 | 0.038 | 0,052 |
| Weight loss/reduced appetite | 39453 | 36497 | 0.174 | 0.077 | 0.025 | 0,045 |
| Feeling worthless | 61757 | 43570 | 0.109 | 0.072 | 0.133 | 0,146 |
| Weight gain/increased appetite | 22612 | 36489 | 0.105 | 0.054 | 0.052 | 0,064 |
| Weight gain, clinical population | 7902 | 13167 | -0.031 | 0.102 | 0.762 | 0,762 |
